# Spatio-temporal spread of COVID-19: Comparison of the inhomogeneous SEPIR model and data from South Carolina

**DOI:** 10.1101/2021.08.15.21262074

**Authors:** Yoav Tsori, Rony Granek

## Abstract

During the COVID-19 pandemic authorities have been striving to obtain reliable predictions for the spreading dynamics of the disease. We recently developed a multi-”sub-populations” (multi-compartments: susceptible, exposed, pre-symptomatic, infectious, recovered) model, that accounts for the spatial in-homogeneous spreading of the infection and shown, for a variety of examples, how the epidemic curves are highly sensitive to location of epicenters, non-uniform population density, and local restrictions. In the present work we test our model against real-life data from South Carolina during the period May 22 to July 22 (2020). During this period, minimal restrictions have been employed, which allowed us to assume that the local basic reproduction number is constant in time. We account for the non-uniform population density in South Carolina using data from NASA, and predict the evolution of infection heat-maps during the studied period. Comparing the predicted heat-maps with those observed, we find high qualitative resemblance. Moreover, the Pearson’s correlation coefficient is relatively high thus validating our model against real-world data. We conclude that the model accounts for the major effects controlling spatial in-homogeneous spreading of the disease. Inclusion of additional sub-populations (compartments), in the spirit of several recently developed models for COVID-19, can be easily performed within our mathematical framework.

## Introduction

Infectious disease spreading models are largely based on the assumption of perfect and continuous “mixing”, similar to the one used to describe the kinetics of spatially-uniform chemical reactions. In particular, the well-known susceptible-exposed-infectious-recovered (SEIR) model, builds on this homogeneous-mixing assumption. Computational tools that include factors such as long range human mobility have been developed to account for pandemic spread on a global scale [1–4]. Some extensions of SEIR-like [5–9] and other models [10–18] that account for spatial variability divided the population to different sub-populations (where the term “sub-population” refers to people under a certain stage of the disease, otherwise termed “compartment”) [19]. Diffusive spreading have been used in numerous models in various levels [20, 21]. To mimic the heterogeneity of human behavior more realistically, recent extensions employed diffusion processes of the sub-populations that are limited to contact networks [22]. However, the application of diffusion process for human population is unrealistic due to its tendency to spread all populations to uniformity (be it in real space or on contact networks). Moreover, most simple diffusive models do not include spatially-dependent population density or a spatial dependence of infection spreading parameters, which are required to model geographically local quarantine. Thus, implementation of such dependencies using homogeneous models requires a division of the geographic region into multiple number of patches [22].

Recently, we presented a general mathematical framework for epidemiological models that can treat the spatial spreading of an epidemic [23]. The framework accounts for possible spatial in-homogeneity of all associated “compartments”, which we termed “sub-populations”, such as the infectious and the susceptible sub-populations. To describe the spatial spreading of COVID-19, we have used the general framework in a 5-compartment model: susceptible, exposed, pre-symptomatic, infectious, and recovered (SEPIR). We used the SEPIR model, with parameters typical for COVID-19, to examine several scenarios of COVID-19 spreading, including the effect of localized lockdowns. However, the validation of our model against the COVID-19 spatial spreading in a specific country is still missing.

Vaccination against COVID-19 is now ongoing in many countries, despite complications associated with shortage of supply, anti-vaccination movements, and vaccination program during a rapidly evolving epidemic. Most strategies use age group and risk factors prioritization, ignoring the density variation of susceptible *vs*. recovered sub-populations. However, under an outbreak it is reasonable to vaccinate first in regions where the outbreak is expected to be stronger. Our in-homogeneous SEPIR model can easily be generalized to predict the outcome of vaccination strategies that involve such variation.

In this publication we wish to validate the in-homogeneous SEPIR model against real world data. High spatial resolution data are difficult to obtain due to issues of personal confidentiality [24]. We have chosen the state of South Carolina which made publicly available COVID-19 infection data and heat-maps with reasonable resolution. Information for the density variation across the country is also readily obtained from NASA public resources. As we have shown [23], these data are critical for the prediction of realistic infection heat-maps and for testing them against those obtained in real-life.

### Inhomogeneous SEPIR model

We briefly review the key features of the in-homogeneous SEPIR model, described in detail in Ref. [23]. To model the spread of COVID-19 and other epidemics, the population is divided into five compartments, termed as “sub-populations”: susceptible, exposed, pre-symptomatic, infectious (that are symptomatic), and recovered. In this generalizations of the SEIR model, the basic chain of infection dynamics is as follows. Healthy individuals, and those that are not immune, are initially susceptible. They can become exposed when in contact with pre-symptomatic (that are also infectious) or infectious-symptomatic people. These individuals stay in the exposed stage for a certain period of time 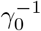 during which they are not infectious to others. After this initial incubation period, they become pre-symptomatic and also infectious to healthy people. They stay as pre-symptomatic for another period of time, denoted by 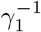, after which they become symptomatic, remaining infectious. This sickness stage lasts for a period of 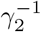 time. At the end of this period, these individuals become recovered and also immune. According to the literature (concerning here only with the parent, wild-type, SARS-COV-2 virus), the mean time for the appearance of symptoms is 5 days, setting 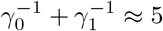 days. It is accepted that people become infectious about 3 days before appearance of symptoms, that is, 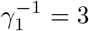 days, and hence 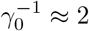 days [25]. The time constants 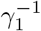 and 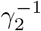, describing the transitions from pre-symptomatic (infectious) to symptomatic (infectious), and from symptomatic-infectious to recovered, respectively, must obey 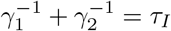, where *τ*_*I*_≈ 16.6 is the average infectious period. It follows that 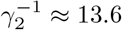 days.

The most unique feature of our model is its ability to account naturally for spatial in-homogeneity. The geographical area under consideration is divided into a lattice that can be square or hexagonal, with inter-node spacing *δ*. At each node, the sum of the five sub-populations equals to the nodal population number (varying from node to node), i.e. the total number of people living in the area corresponding to that node.

The spatial dependence of the number of people in each sub-population is accounted for by inclusion of infection kinetics between neighboring nodes (“interactions”). On a scale much longer than the inter-node spacing, all densities become continuous. In this limit, inter-node interactions give rise to diffusion-like infection terms in the rate equations. After proper rescaling, the model equations can be written as [23]

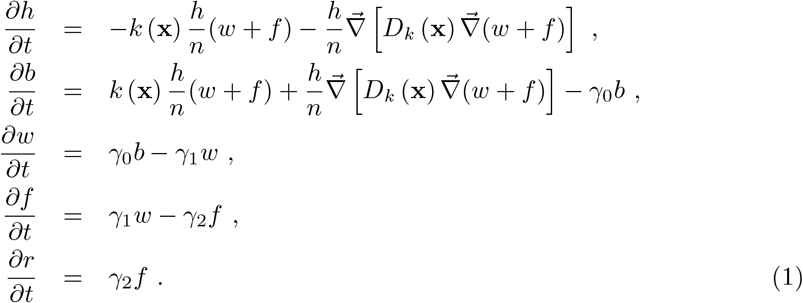

The variables *h, b, w, f*, and *r* denote the number area density of susceptible (healthy), exposed, pre-symptomatic, symptomatic, and recovered sub-populations, respectively. They are dimensionless due to scaling by the average population density in the region under study. These variables depend on the spatial location **x** (scaled by *δ*). Their sum obeys

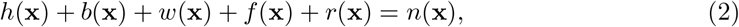

where *n*(**x**) is the total (fixed) population density at location **x**. We emphasize that the above rate equations describe diffusion of the epidemic and *not* of people; at any point **x** the sum *h*(**x**) + *b*(**x**) + *w*(**x**) + *f* (**x**) + *r*(**x**) remains constant in time.

The spatially-dependent parameters *k*(**x**) and *D*_*k*_(**x**) originate both from infections within each node (influencing *k* alone), and from infections between neighboring nodes (influencing both *k* and *D*_*k*_) [23, 26]. *k* describes the rate at which susceptible people (*h*) become exposed (*b*) when they meet asymptomatic (*w*) or symptomatic (*f*) people. It is given by *k* = *R*_0_*/τ*_*I*_, where *R*_0_ is the well known basic reproductive number. The epidemic diffusion coefficient is estimated to be *D*_*k*_≈ *k/*5 for a square lattice. Our model thus consists of five nonlinear partial differential equations in two spatial dimensions. They can be solved numerically with given initial conditions and a given total population density *n*(**x**), as we present next using real-world data from South Carolina.

## Methods

In the years 2020–2021 the COVID-19 epidemic has spread to many places around the world [27]. Here we focus on South Carolina, which has been hit hard by the virus and has high quality data available in the public domain. Specifically, the South Carolina Department of Health and Environmental Control (DHEC) provides areal maps and tabulated data of infected people density with daily resolution [28, 29]. The number of cases is given per ZIP code (sub-county level), and hence the state is divided into ∼ 400 small regions, providing excellent spatial resolution. The second source of data is the “Covidtracking” website [30], including various whole state (i.e. spatially integrated) quantities, such as the number of active cases, daily new cases, and accumulated cases. The third data source we used is NASA’s Socioeconomic Data and Applications Center (SEDAC) [31], from which we obtained the South Carolina population density *n*(**x**).

We restrict ourselves to the time window between May 22, 2020 (initial time *t* = 0) and July 22, 2020; see the heat-maps in Fig. 2 for the reported initial (May 22, 2020) and final (July 21, 2020) COVID-19 infections. During this period, no major South Carolina government orders were issued [32]. After a long period of restrictions, a series of “opening” executive orders commenced on April 21 and ended with the following four relevant opening orders: (i) on May 8, (ii) on May 11, (iii) on May 22 [33], and (iv) on June 12 [34]. Whereas the orders on May 22 and June 12 can possibly have some influence on the transmission of COVID-19 during the period of study May 22–July 22 (noting that infection data may lag the actual infection events up to 7-10 days), we believe they are of minor consequence. The next state government executive order is on July 10 and is of restriction type; Due to the data lag, it may affect the published infection data only at the very last few days of the studied period. Therefore, we assume that *R*_0_remained almost constant during the studied period and time-dependent adjustments are not required. Indeed, this two months period is a strong test of our model since common predictions using homogeneous models usually require parameter adjustments after a few weeks at most. In addition, human mobility studies show that the mean travel distance during this period was rather short, mostly much below 30 miles [35]. Such conditions are ideally suited for our model, and suggest an upper bound estimate for the node size, *δ* = 30 miles, and that node-to-node infections are dominant.

To obtain the population density *n*(**x**) we read the NASA SEDAC image [31, 36] and cropped it to the relevant South Carolina region, see Fig. 1. The image was converted to shades of gray (bright meaning high population density), then scaled and rotated by 2 degrees to fit the images from DHEC. Each pixel corresponds to a unique location **x** in our model. We obtained the pixel area 3.9 km^2^ by counting the number of pixels within the South Carolina borders and dividing by the state area, 82, 930 km^2^. The two-dimensional matrix *n*(**x**) was calculated in by dividing the pixel brightness at each location **x**, varying between 0 and 1, by the average pixel brightness over the area of the state.

**Fig 1.**
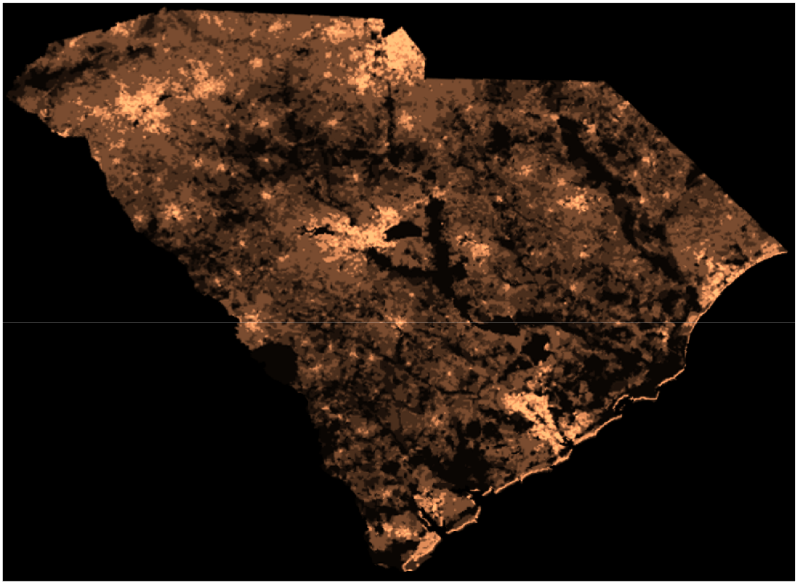
Population density of South Carolina from NASA’s SEDAC. Bright spots correspond to highly populated regions. Population outsides of the state’s limit was ignored (large dark areas).

Infection data were obtained from the South Carolina DHEC in the form of tables with the number of confirmed cumulated cases for each zip code, defined as *c*. For the initial conditions, we assume that the global ratio of active infections to cumulated infections, *q* = (*W* + *F*)*/*(*F* + *R*), holds also locally, i.e. it is equal to (*w* + *f*)*/*(*f* + *r*). The global ratio was taken from Covidtracking.com [30], and on May 22, 2020, it was equal to *q* = 3304*/*9766. Since, by definition, *c* = *w* + *f* + *r*, we obtain for the local quantities *r* = *c*(1 −*q*) and *f* + *w* = *cq*. Further, to find separately *f* and *w*, we assumed that each of these variables is proportional to the fraction of time of the corresponding disease stage out of the total infection time, i.e. 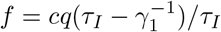 and 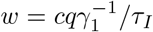. For *h* and *b* at *t* = 0 we took *b* = 0 and *h* = *n* − *f* − *w* − *r*. Lastly, to obtain all model variables on the resolution of the NASA’s SEDAC population density map, Fig. 1, model variables were multiplied by the ratio of local to global population densities, *n*(**x**). For example, this converts the two panels in Fig. 2 to the respective panels in Figs. 3 (top-left) and 4 (bottom-left).

**Fig 2.**
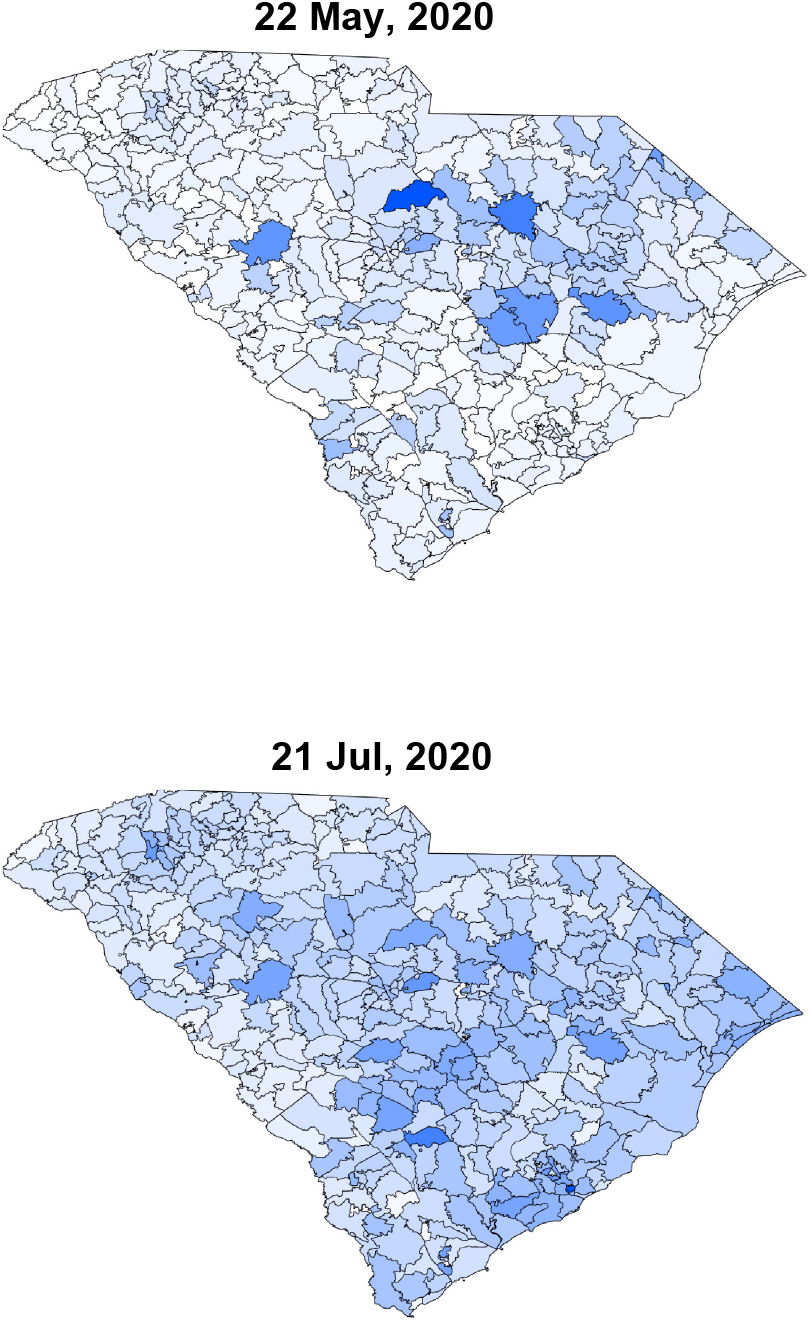
Infection maps taken from the South Carolina DHEC. The top figure corresponds to the initial time of the simulation (*t* = 0), while the bottom, two months later, is the last time. The top figure serves as the initial condition for the simulations.

**Fig 3.**
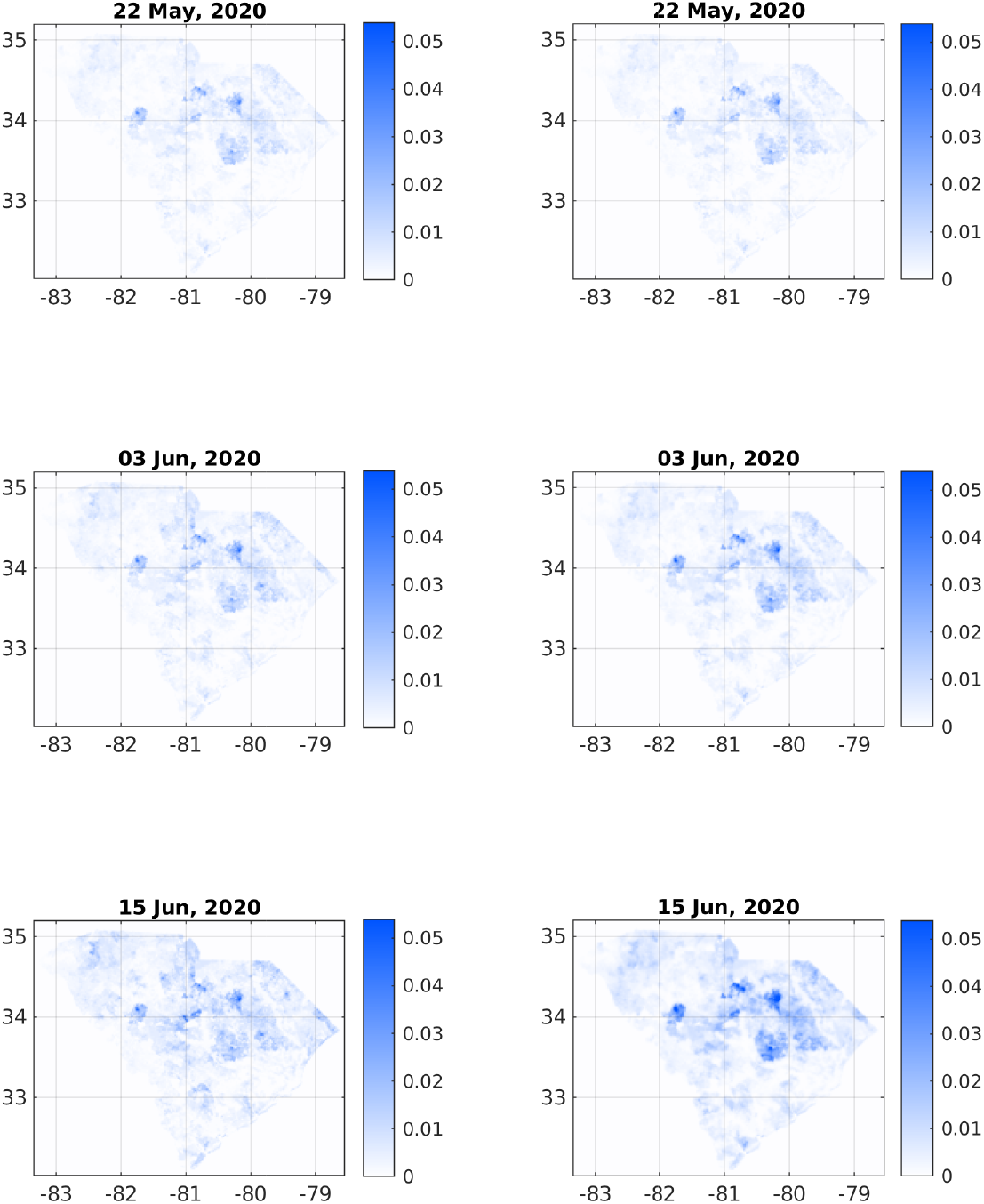
Comparison of infection dynamics between the data from the DHEC (left column) and the theoretical model (right column). In both columns, images correspond to the sum *f*(**x**) + *w*(**x**) + *r*(**x**). The heading for each panel shows the date, starting from May 22, 2020, corresponding to relative times *t* = 0, *t* = 12, and *t* = 24 days. See later times in Fig. 4.

It is estimated that the actual number of infected people in the community, during the studied period, is about 5-15 times larger than the number of positive RT-PCR tests, as implied by several seroprevalence surveys of antibodies to SARS-CoV-2 around the US [37–39] and in Israel [40]. In South Carolina, the ratio between the estimated and known infected numbers is approximately 5 [28]. Accordingly, to estimate the actual number of infected population, the observed density was multiplied by a factor 5. To make a valid comparison with the reported positive RT-PCR number of infected people, we divided back the results of the simulations by 5.

Finally, we used shape-files taken from the United States Census Bureau [41] to allocate the number of cases to the proper geographical location, and digitized the cases into the same grid (matrix) as the one used for the population density. In Fig. 2 (top) we depict the cases heat-map as a visual presentation of the initial conditions used in the simulations.

To solve the set of coupled rate equations Eqs. (1), we used the same two-dimensional grid of the DHEC data. The gradient and divergence operators were discretized according to standard finite difference formulations. To propagate the equations in time we used an explicit scheme with a sufficiently small time differential.

To calculate the Pearson’s correlation coefficient (CC) between observed and predicted heat-maps [42] we defined *I*_data_≡ *I*_data_(**x**_*j*_) as the infected matrix of the sum *f* + *r* + *w* taken from the DHEC data, and *I*_sim_≡ *I*_sim_(**x**_*j*_) as the sum *f* + *r* + *w* from the simulations. The Pearson’s CC is then defined as

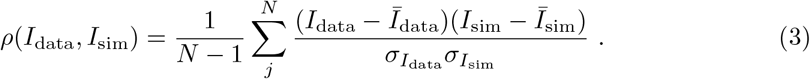

The index *j* runs over all pixels (**x** coordinates), 0 ≤*j* ≤*N*, and *N* is the total number of pixels in the images. *Ī*_data_and *Ī*_sim_are the averages of *I*_data_and *I*_sim_, respectively, and 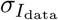 and 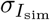 are their standard deviations.

## Results

Figures 3 and 4 depict a comparison between six pairs of reported *vs* predicted South Carolina heat-maps from the period under study. May 22, 2020 serves as the zero time for simulation and therefore the two top images are the same. The visual similarity between the South Carolina heat-maps and the simulations for the subsequent dates, including the very late date of July 21, is evident.

**Fig 4.**
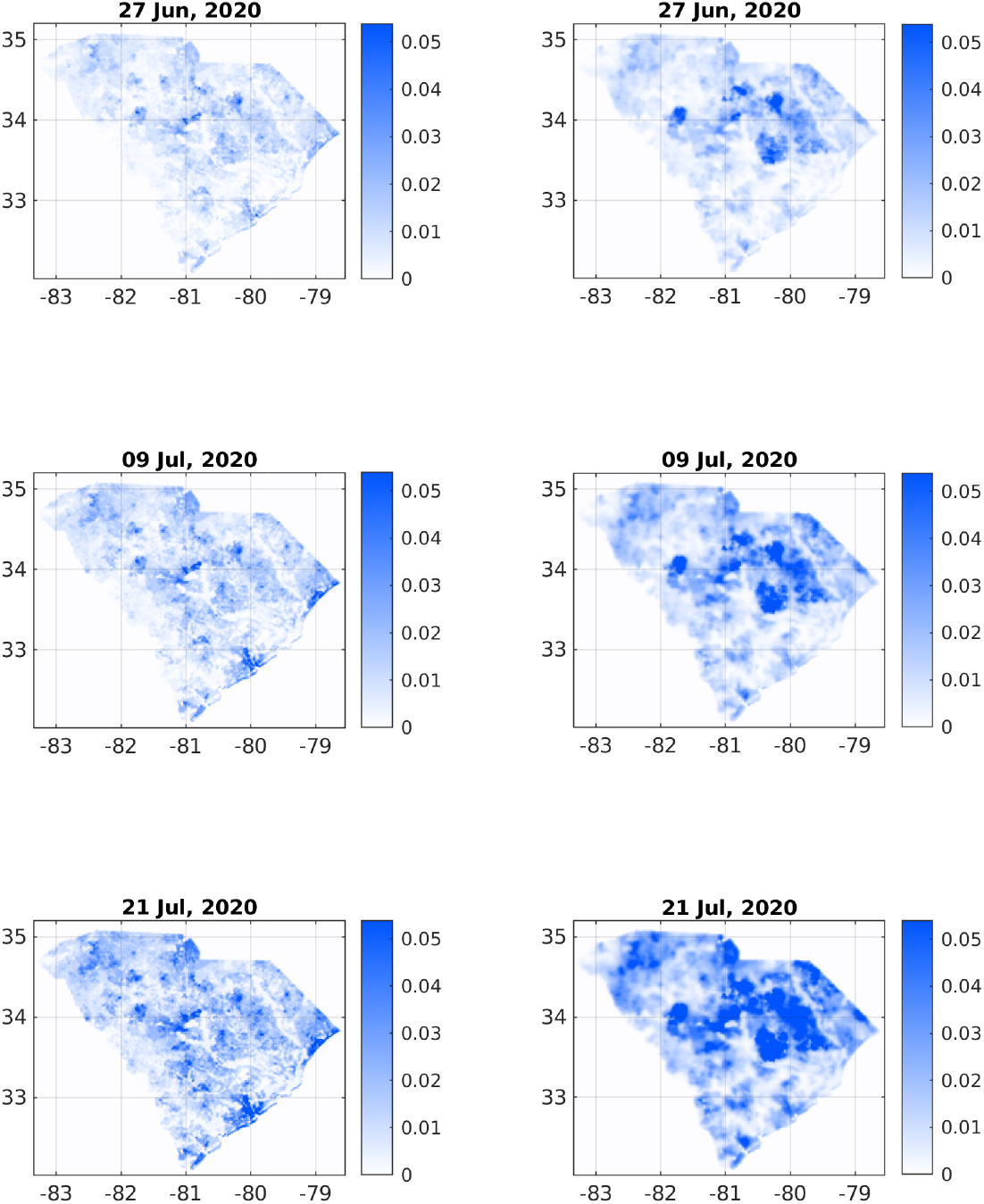
Comparison of infection dynamics – continuation of Fig. 3 for relative times *t* = 36, *t* = 48, and *t* = 60 (dates June 27, July 9, and July 21, respectively).

To make this comparison more quantitative, we calculated the Pearson’s CC as defined in Eq. (3). Table 2 shows the Pearson’s CC values, *ρ*(*I*_data_, *I*_sim_), for all the six dates corresponding to the heat-maps in Figs. 3 and 4. Not surprisingly, the CC value is 1 for the initial conditions (top pair panels in Fig. 3). For the subsequent dates, the CC values are slowly descending – which is quite reasonable as errors accumulate in time – yet remain close to unity. For July 21 (2020) we find *ρ*(*I*_data_, *I*_sim_) ≈ 0.77, which is still quite high, recalling that we ran our simulation for 60 days without any intermediate tweaking of the model parameters. Figure 5 (a) supplements Table I to all dates in the studied period, using a fitted interpolation formula.

**Table I.** Summary of variables of the model [Eqs. (1) and (2)] and their respective time scales.

| Variable name | incubation time | details |
| --- | --- | --- |
| h |  | density of susceptible (healthy) |
| b | $\gamma_0^{-1} = 2$ days | density of exposed |
| w | $\gamma_1^{-1} = 3$ days | density of pre-symptomatic and infectious |
| f | $\gamma_2^{-1} = 13.6$ days | density of symptomatic and infectious |
| r |  | density of recovered |
| n |  | density of total population |

**Table II.**
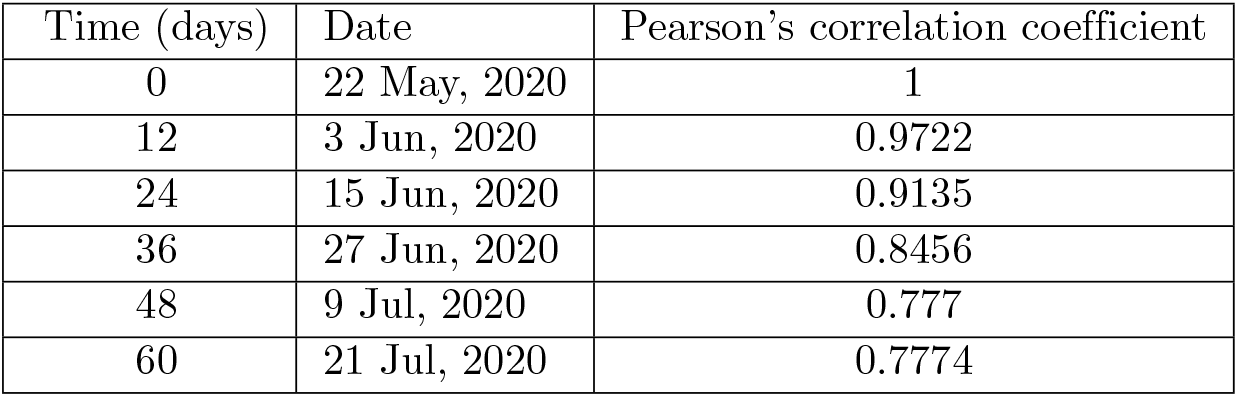
Pearson’s correlation coefficient between the reported (left column) and simulated (right column) cases shown in Figs. 3 and 4. The correlation coefficient decays in time but is still high even after 60 days.

**Fig 5.**
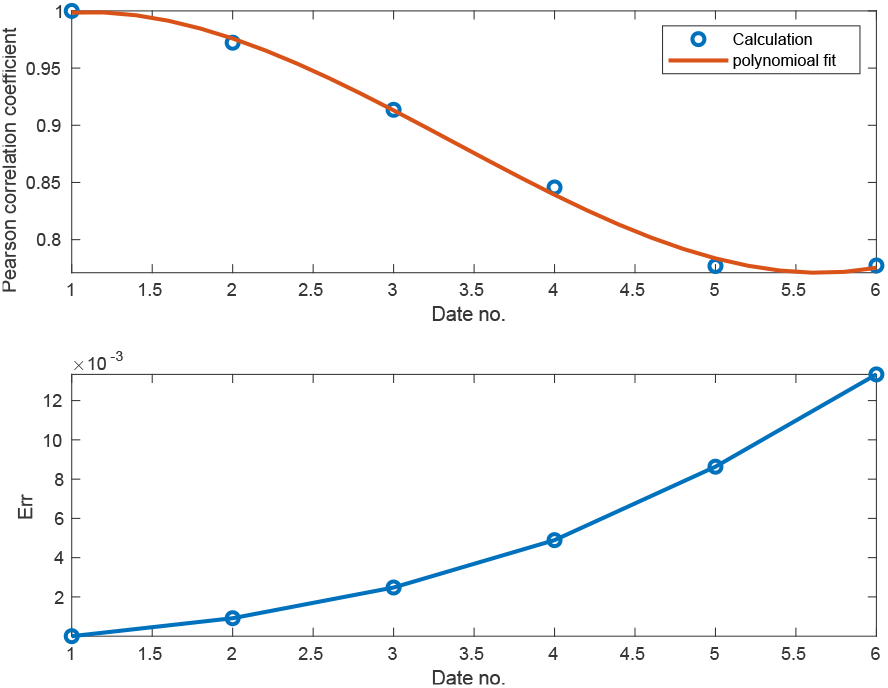
(a) Pearson correlation coefficient between simulation and observation from Eq. (3) vs time. A high value of correlation coefficient means good agreement. By definition of the initial conditions, the correlation starts from the maximum value, 1. It then monotonously decays with time. (b) Estimated root-mean-square error *Err* as defined in Eq. (4). The error starts from zero and increases with time.

To further confirm the similarity between predicted and observed heat-maps, we performed a root mean square analysis, see Fig. 5 (b). The variable on the vertical axis, *Err*, is defined as follows:

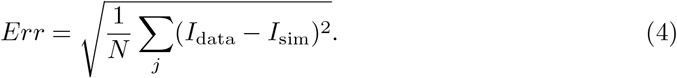

*Err* gives an estimate of the overall “error” in our predictions. Perfect match should ideally yield *Err* = 0. Notably, the error here is below 0.01 (except at the very last days of the simulation), which is five times smaller than the maximum of *I*_data_, indicating small differences between prediction and observation.

Figure 6 compares the total (integrated) number of cases predicted by the theory against the actual reported values. The dashed red curve is the number of cases [30] while the blue curve is the theory. Time *t* = 0 corresponds to May 22, 2020; by definition of the initial conditions, the theoretical value is the same as the reported one. The agreement between the simulation and the reported numbers is quite good, even though during the whole 60 days simulation time the basic reproduction rate and other model parameters were held constant. The final simulated number of cases, 49, 085, is 6% above the reported number, 46, 275.

**Fig 6.**
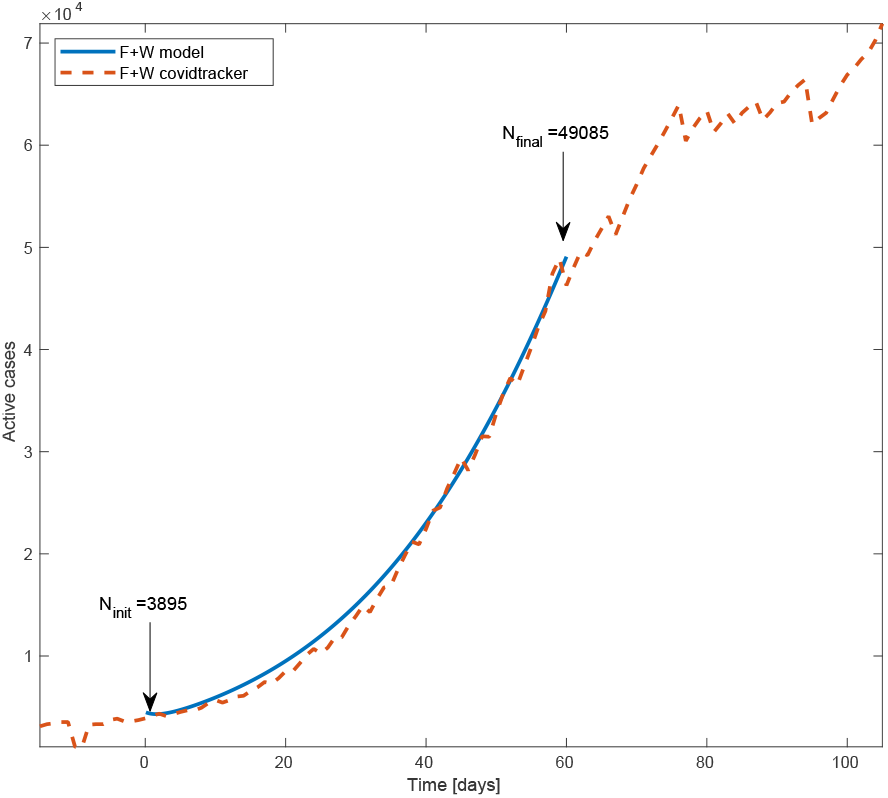
Total number of active cases (’*F*’+’*W*’) in South Carolina vs time. Blue line is the model simulation and red dashed curve is the number provided by Covidtracking. The model input is the total number of active cases at time *t* = 0 (May 22, 2020), 3895. The final number of cases of the model is 49, 085, about 6% above the reported number, 46, 275. We assumed the basic reproductive rate *R*_0_= 1.9 to be constant throughout the whole inspected time.

## Conclusions

In this paper we have tested the in-homogeneous SEPIR model against spatial data from South Carolina and found remarkable agreement of the infection heat-maps. The visual agreement between the model and the data is confirmed by the large Pearson’s correlation coefficient. The implementation of the current approach to the COVID-19 pandemic in other parts of the world requires extended and high resolution data of the geographic spread of the pandemic, to serve as the models’ initial conditions for the five sub-populations. Also required by the model is a high resolution data of the geographic population density.

The model’s general mathematical framework can be easily implemented in essentially all “multi-compartment” (i.e. several sub-population) models developed so far for the COVID-19 pandemic [13, 17, 43]. It may be also used to predict the effect of vaccination, by adjusting the local fraction of susceptible sub-population *h*(**x**)*/n*(**x**), thereby helping to improve vaccination strategies. We hope that further use and development of our approach will assist fighting the COVID-19 and other pandemics. Further improvements of our model are currently underway.

## Supporting information

Supplemental movie of simulated heat-maps

Supplemental movie of observed heat-maps

## Data Availability

1) All simulations data will be provided uppon request.
2) The sources of data used for comparsion with the simulations are properly referenced in the manuscript.

## Acknowledgments

We are grateful to Ariel Kushmaro for insightful discussions.

